# Data-efficient machine learning for the detection of duodenal neoplasia: a multi-centre study

**DOI:** 10.64898/2026.10.08.26365053

**Authors:** Flavio Di Lisio, Elizabeth J. Soilleux, Florian Jaeckle

## Abstract

**Background:** Machine Learning (ML) has shown promise in histopathological diagnosis, but rare tumours, like duodenal epithelial neoplasms, are challenging because large, labelled datasets are difficult to assemble. We propose a data-efficient ML pipeline for detecting duodenal epithelial neoplasia in whole-slide images.

**Methods:** We assembled a five-hospital dataset of haematoxylin and eosin–stained duodenal biopsy whole slide images (WSIs) at 10x magnification, comprising 8,027 normal and 401 neoplastic biopsies, including adenomas, carcinomas and neuroendocrine tumours. WSIs were divided into patches and encoded using the Hibou-B foundation model. We compared global standardisation with unsupervised site-specific standardisation of patch embeddings. Our pipeline combined principal component analysis, a Gaussian mixture model and a Random Forest classifier. We assessed generalisability using leave-one-hospital-out cross-validation across four hospitals and on an independent test set from a fifth hospital.

**Results:** In leave-one-hospital-out cross-validation, the pipeline achieved a mean accuracy of 90%, sensitivity of 96%, specificity of 83% and an AUC of 0.96. On the external test hospital, it achieved 94% accuracy, 93% sensitivity, 95% specificity, and AUC of 0.97. Site-specific standardisation improved mean cross-validation accuracy by 20 percentage points compared with global standardisation, while the Gaussian mixture pipeline improved accuracy by 12 percentage points compared with mean pooling of patch embeddings.

**Conclusions:** We present a data-efficient ML approach for detecting duodenal epithelial neoplasia from WSIs. Strong performance was maintained across hospitals and on an independent external test cohort despite the limited number of neoplastic training cases, supporting the feasibility of foundation-model-based approaches for rare histopathological diagnoses.

## Introduction

The rise of digital pathology has created new opportunities for the application of machine learning (ML) to histopathological diagnosis. ML-based tools have the potential to reduce inter-observer variability^1^, support pathologists facing high workloads^2^ and extend high-quality diagnostic capabilities to centres with limited access to specialist pathologists^3^.

While many developments in computational pathology have focused on common malignancies such as lung, breast, prostate, and colorectal cancer^4,5^, duodenal epithelial neoplasia remains, to the best of our knowledge, entirely unexplored. Duodenal epithelial neoplasms, including adenomas, adenocarcinomas, and neuroendocrine tumours (NETs), are rare: they account for approximately 2% of all small intestinal tumours^6^. Furthermore, our large single-centre audit^7^ of 6245 duodenal biopsies showed that adenomas, carcinomas and NETs combined accounted for only 2.37% of all duodenal biopsies.

Early detection of duodenal epithelial neoplasms is clinically important. Duodenal adenomas carry a malignant potential, while duodenal adenocarcinomas are associated with a 5-year overall survival rate of approximately 30-40%^8^. Duodenal NETs, although often indolent, may require surgery or surveillance depending on tumour size, depth of invasion, and grade^9^.

Developing ML models that generalise across different centres typically requires large and diverse multicentre datasets. This is particularly challenging for rare conditions, such as duodenal epithelial neoplasia, where case numbers are inherently limited. Furthermore, inter-centre variability that stems from differences in tissue preparation techniques, staining protocols and digital scanning systems^10,11^ may hinder training and introduce spurious centre-specific signals, potentially causing the models to overfit to centre-specific technical artefacts rather than learning diagnostically relevant features. Achieving robust performance across institutions therefore remains one of the major barriers to clinical deployment of computational pathology systems. These challenges make duodenal neoplasia an attractive application for data-efficient approaches that can leverage prior knowledge learned from large histopathology datasets. In this context, the recent development of foundation models pretrained on a large and diverse histopathology dataset represents a significant advance for computational pathology. By leveraging representations learned during large-scale pretraining, these models can extract rich morphological features that can be used by downstream classifiers without requiring large amounts of labelled data that would otherwise be necessary to train a model from scratch. We employ Hibou-B^12^, a vision transformer-based foundation model pretrained on a large and diverse collection of pathology images, including gastrointestinal biopsies, to extract patch-level embeddings from haematoxylin and eosin (H&E)-stained duodenal biopsy whole-slide images (WSIs).

In this work, we employ a Gaussian Mixture Model (GMM) based pipeline for detecting duodenal epithelial neoplasms using patch-level embeddings extracted by Hibou-B. Biopsy-level representation vectors are constructed using a GMM and classified using a Random Forest model. We investigate whether site-specific standardisation of patch embeddings improves cross-site generalisation compared with global standardisation. We assess model performance using leave-one-hospital-out cross-validation across four hospitals and further evaluate the pipeline on an independent external test set from a previously unseen fifth hospital using a scanner from a different manufacturer. To our knowledge, this is the first study to develop and evaluate an ML pipeline for the detection of duodenal epithelial neoplasms from whole-slide images.

## Methods

### DATASET

We collected a training dataset consisting of a total of 6,660 WSIs from 6,162 patients, including 6,317 normal duodenal biopsies and 272 adenomas, 60 carcinomas and 11 NETs. The biopsy WSIs were obtained from 4 different hospitals (Addenbrooke’s Hospital, Cambridge; Glasgow Queen Elizabeth Hospital; University Hospital of North Tees; and the University Hospitals of Coventry & Warwickshire (UHCW)) (Table 1). Each hospital used digital scanners from different manufacturers. WSIs from Addenbrooke’s Hospital comprised two cohorts acquired in different years using different scanners: cohort 1 included 2,783 WSIs, consisting of 2,632 normal and 151 neoplastic (130 adenomas, 13 carcinomas and 8 NETs). Cohort 2 included 996 WSIs, consisting of 938 normal and 58 neoplastic (29 adenomas, 27 carcinomas and 2 NETs).

**Table 1.** Our training dataset includes 6,660 WSIs from four hospitals and our test set 1768 WSIs from a fifth hospital.

|  | Train |  |  |  | Test |  |  |  |
| --- | --- | --- | --- | --- | --- | --- | --- | --- |
|  | Normal | Adenoma | Carcinoma | NET | Normal | Adenoma | Carcinoma | NET |
| Addenbrooke's Hospital, Cambridge | 3570 | 159 | 40 | 10 | — | — | — | — |
| Queen Elizabeth Hospital, Glasgow | 540 | 12 | 2 | 0 | — | — | — | — |
| University Hospital of North Tees | 1722 | 28 | 5 | 0 | — | — | — | — |
| University Hospitals of Coventry and Warwickshire | 485 | 73 | 13 | 1 | — | — | — | — |
| University Hospital of Southampton | — | — | — | — | 1710 | 41 | 7 | 10 |
| Male | 2641 | 148 | 28 | 4 | 774 | 25 | 2 | 5 |
| Female | 3676 | 124 | 32 | 7 | 936 | 16 | 5 | 5 |
| 0-9 | 40 | 0 | 0 | 0 | 238 | 0 | 0 | 0 |
| 10-19 | 132 | 0 | 0 | 0 | 746 | 0 | 0 | 0 |
| 20-29 | 334 | 2 | 1 | 0 | 69 | 0 | 0 | 0 |
| 30-39 | 460 | 8 | 0 | 0 | 67 | 0 | 0 | 0 |
| 40-49 | 742 | 16 | 1 | 2 | 98 | 11 | 0 | 6 |
| 50-59 | 1028 | 22 | 7 | 1 | 99 | 5 | 1 | 3 |
| 60-69 | 1288 | 62 | 12 | 3 | 159 | 17 | 3 | 0 |
| 70-79 | 1465 | 116 | 29 | 4 | 165 | 6 | 0 | 1 |
| 80-89 | 776 | 40 | 9 | 1 | 65 | 2 | 3 | 0 |
| 90-99 | 52 | 6 | 1 | 0 | 4 | 0 | 0 | 0 |

Our test dataset included a total of 1,768 WSIs from the University Hospital of Southampton from 1,075 patients, including 1,710 normal duodenal biopsies, 41 adenomas, 7 carcinomas and 10 NETs.

WSIs in the training or test set that had fewer than 100 patches (as defined below) were excluded from analysis (125 normal and 0 neoplasm cases). All WSIs and accompanying fully anonymised patient data were obtained with Health Research Authority (HRA) Approval (reference: 335851, issued on 18 December 2023), which stated that specific patient consent was not required for the study.

### WHOLE-SLIDE IMAGE PREPROCESSING

We first selected tissue-containing regions and removed unwanted artefacts using Schreiber’s method^13^ as described in previous studies^14,15^. From these regions, we extracted 256 × 256-pixel patches at 10x magnification (1µm per pixel) using a sliding-window approach with a stride of 128 pixels. We normalised pixel intensities using Macenko’s method, which has been shown to improve performance and increase inter-hospital generalisability^11,16,17^. Each patch was converted into a 768-dimensional embedding vector using the Hibou-B foundation model^12^.

### MODELS UTILISED

We developed a GMM-based pipeline implemented in Python (version 3.10.9) using the Scikit-learn library (version 1.6.1)^18^. We first performed a standardisation step. For each hospital *H*, we calculated in an unsupervised manner the mean vector 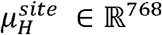 and the standard deviation vector 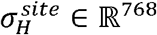 across the embedding vectors of all training patches from hospital *H*. From each biopsy *x*, let *H*_*X*_ denote the hospital from which it comes. Let *x*_*i*_ denote the feature vector extracted by Hibou-B from patch *i* of biopsy *x*. Its site-specific standardised embedding was computed as:

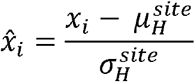

The normalised patch embeddings 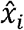 were subsequently reduced to 50 dimensions via Principal Component Analysis (PCA) 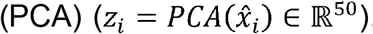, after which a GMM with K = 50 Gaussian components was fitted using the expectation-maximization algorithm on the reduced embeddings from all training patches. In training, we employed diagonal covariance matrices, a maximum number of iterations of 200 and K-means initialisation with 10 random initialisations. For each patch *z*_*i*_, after dimensionality reduction, the GMM computed the posterior probability (assignment probability) *p*(*k*|z_*i*_) of belonging to each of the 50 Gaussian components. Within each leave-one-hospital-out fold, PCA and the GMM were fitted exclusively using patch embeddings from the training hospitals and were subsequently applied without refitting to the hold-out hospital dataset (University Hospital of Southampton). Site-specific standardisation parameters for the hold-out hospital were estimated independently from its unlabelled patch embeddings.

For a biopsy containing *N* patches, we calculated the biopsy-level average assignment probability vector *a* by averaging the GMM posterior probability vectors across patches:

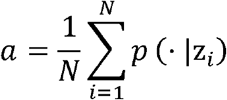

Let 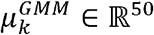 denote the mean vector of Gaussian component *k*, for *k* = 1, …*K*. For each PCA-transformed patch embedding *z*_*i*_, we computed an expected GMM representation *r*_*i*_ ∈ ℝ^50^, as the posterior-probability-weighted average of the component means (using the patch’s assignment probabilities *p*(*k*|*z*_*i*_) as weights):

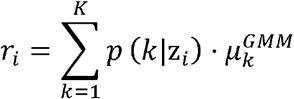

We subtracted these reconstructions from the PCA-transformed patch embeddings, obtaining a residual vector s for each patch, which was L2-normalised.

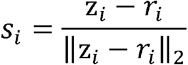

These vectors were then aggregated into a single biopsy-level residual 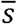 through mean-pooling.

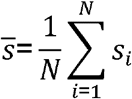

We finally constructed a final biopsy embedding *b* ∈ ℝ^100^ by concatenating 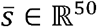 with the average assignment probability vector *a* ∈ ℝ^50^:

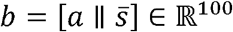

These embeddings were then passed to a final classifier, which was a Random Forest model, with 100 estimators with no maximum tree depth imposed.

### COMPARING GLOBAL AND SITE-SPECIFIC STANDARDISATION

To evaluate the importance of site-specific standardisation, we compared our proposed method with global standardisation, in which the feature-wise mean (μ) and standard deviation (σ) were estimated jointly from all training patch embeddings across all training hospitals. These parameters were fixed and subsequently applied to embeddings from the hold-out site.

To assess the contribution of the GMM-based biopsy representation, we additionally compared our GMM-based pipeline with mean-pooling, in which, after site-specific standardisation, the biopsy representation vector *b*_mean_ ∈ ℝ^768^ was constructed by calculating the mean of its raw patch embeddings:

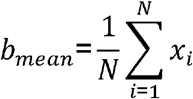

The resulting biopsy-level representation was passed to a Random Forest classifier with identical hyperparameters as the classifier used in the GMM-based pipeline.

### MODEL DEVELOPMENT

We trained our GMM-based pipeline and the mean pooling baseline on the training dataset using leave-one-hospital-out cross-validation. To maintain class balance within each hospital, we included all available epithelial neoplastic biopsy WSIs from each hospital and randomly sampled an equal number of normal biopsy WSIs from the same hospital. Biopsies were treated as independent samples for model training and evaluation. Some patients contributed more than one biopsy; however, because cross-validation was performed by holding out an entire hospital, biopsies from the same patient could not occur in both the training and validation sets. Because all available neoplastic WSIs were used from each hospital, the size of the validation set and of the training set varied between folds. The reported mean is an unweighted average across hospitals. During model training and validation, 100 patch embeddings were randomly sampled from each biopsy. During external testing, all available patch embeddings were used for each biopsy. Each leave-one-hospital-out fold was repeated five times, each time drawing a different random sample of normal biopsy WSIs and patch embeddings. For the test set experiments, we trained our models using data from all four training hospitals, and evaluated them on the test set, which contained 1768 WSIs, including 58 biopsy WSIs containing epithelial neoplasms. Results are reported as mean ± standard deviation across the five repetitions.

### ADDITIONAL INVESTIGATIONS

We performed additional analyses to investigate the information retained in the biopsy and patch representations after preprocessing.

We used Uniform Manifold Approximation and Projection (UMAP) to visualise the biopsy-level representations generated by the GMM pipeline. We compared representations obtained using global and site-specific standardisation and visualised the resulting projections according to both hospital of origin and diagnostic label (normal or neoplasm). For these analyses, we included all available neoplastic biopsy WSIs and an equal number of normal biopsy WSIs randomly sampled from each respective hospital.

To quantify the extent to which site-specific information remained encoded after preprocessing, we assessed whether the hospital of origin could be predicted from the patch representations following global or site-specific standardisation. We used 28 biopsy WSIs from each training hospital, comprising 14 neoplastic and 14 normal biopsy WSIs, randomly selected from all biopsies in each category at each hospital. Fourteen neoplastic biopsy WSIs were used per hospital because this was the maximum available from the hospital with the fewest neoplastic cases. Patches extracted from these biopsy WSIs were used as individual samples for hospital of origin prediction. The experiment was repeated five times, with a different sample of biopsy WSIs selected in each repetition.

In order to assess the applicability of site-specific standardisation, we conducted an ablation study to investigate the number of biopsy WSIs needed to estimate its parameters in a previously unseen hospital (Supplementary Materials A). We additionally assessed whether the learned representations retained information about patient sex and age (Supplementary Materials B). To assess which type of neoplasm was the hardest to classify, on our independent test set, we calculated the AUC for distinguishing normal biopsies from adenomas, carcinomas, and NETs separately (Supplementary Materials C). We also conducted an ablation study to assess the impact of the patch sampling strategy on model performance (Supplementary Materials D). Specifically, we compared using all available patch embeddings for each sampled biopsy with randomly sampling 100 patch embeddings for each sampled biopsy. We also evaluated the performance of our GMM-based pipeline with site-specific standardisation at the patient level (Supplementary Materials E). Full details and results of these analyses are provided in the Supplementary Materials.

## Results

### CROSS-VALIDATION PERFORMANCE

In leave-one-hospital-out cross-validation across four hospitals, the model with site-specific standardisation achieved a mean accuracy of 90% ± 4.2%, a specificity of 83% ± 6.5%, sensitivity of 96% ± 3.8%, and AUC of 0.96 ± 0.025 (Table 2). The performance of the model was consistent across hospitals, with an accuracy ranging from 85% to 95%, sensitivity from 93% to 100%, specificity from 77% to 92%, and AUC from 0.93 to 0.99. These ranges should be interpreted with caution, as the hospitals contributed different numbers of WSIs (Table 1). In particular, estimates for the smallest cohorts (Glasgow and North Tees) are subject to greater sampling uncertainty.

**Table 2.** Performance of the GMM-based pipeline using site-specific standardisation (GMM with ss), the GMM-based pipeline using global standardisation (GMM with gs) and the mean pooling baseline (baseline) with site-specific standardisation applied. Accuracy, sensitivity, specificity, and area under the receiver operating characteristic curve (AUC) are reported as mean ± standard deviation across five repetitions with different random seeds. Neoplasia was considered the positive class. In the Mean column, the average metrics across hospitals ± the standard deviations across folds are reported. For each metric, the bolded value indicates the best mean performance among the three pipelines.

| Hospital used as validation | Addenbrooke's Hospital, Cambridge | Queen Elizabeth Hospital, Glasgow | University Hospital of North Tees | University Hospitals of Coventry and Warwickshire | Mean |
| --- | --- | --- | --- | --- | --- |
| GMM with ss, accuracy | 88% ± 1.6% | 90% ± 4.8% | 95% ± 0.9% | 85% ± 4.8% | **90% ± 4.2%** |
| GMM with gs, accuracy | 59% ± 2.3% | 70% ± 10.4% | 70% ± 11.9% | 79% ± 7.4% | 70% ± 8.2% |
| baseline accuracy | 65% ± 2.1% | 80% ± 9.3% | 88% ± 3.5% | 77% ± 2.7% | 78% ± 9.5% |
| GMM with ss, sensitivity | 93% ± 2.9% | 100% ± 0% | 99% ± 1.3% | 93% ± 2.1% | 96% ± 3.8% |
| GMM with gs, sensitivity | 97% ± 1.7% | 100% ± 0% | 98% ± 2.5% | 92% ± 1.5% | **97% ± 3.4%** |
| baseline sensitivity | 96% ± 1.2% | 100% ± 0% | 85% ± 4% | 86% ± 1.6% | 92% ± 7.4% |
| GMM with ss, specificity | 84% ± 2.2% | 80% ± 9.3% | 92% ± 1.6% | 77% ± 8.7% | **83% ± 6.5%** |
| GMM with gs, specificity | 22% ± 4.4% | 40% ± 20.6% | 42% ± 25.2% | 69% ± 15.2% | 43% ± 19.4% |
| baseline specificity | 34% ± 3% | 60% ± 18.6% | 92% ± 3.9% | 70% ± 6.2% | 64% ± 24.1% |
| GMM with ss, AUC | 0.96 ± 0.007 | 0.93 ± 0.088 | 0.99 ± 0.014 | 0.95 ± 0.02 | **0.96 ± 0.025** |
| GMM with gs, AUC | 0.82 ± 0.034 | 0.88 ± 0.079 | 0.95 ± 0.027 | 0.92 ± 0.03 | 0.89 ± 0.056 |
| baseline AUC | 0.88 ± 0.012 | 0.88 ± 0.055 | 0.94 ± 0.017 | 0.92 ± 0.02 | 0.91 ± 0.03 |

**Table 3.** Results of our ablation experiment investigating how performance varies with the number of biopsies used to estimate the parameters for site-specific standardisation on the validation set. Each experiment was repeated five times, results are reported as mean ± standard deviation.

| Hospital used as validation | 5% used, AUC | 10% used, AUC | 20% used, AUC | 30% used, AUC | 40% used, AUC | 50% used, AUC | 100% used, AUC |
| --- | --- | --- | --- | --- | --- | --- | --- |
| Addenbrooke's Hospital, Cambridge | 0.96 ± 0.013 | 0.97 ± 0.003 | 0.97 ± 0.004 | 0.97 ± 0.006 | 0.97 ± 0.006 | 0.97 ± 0.01 | 0.97 ± 0.005 |
| Queen Elizabeth Hospital, Glasgow | 0.93 ± 0.097 | 0.93 ± 0.074 | 0.92 ± 0.096 | 0.89 ± 0.092 | 0.92 ± 0.081 | 0.9 ± 0.063 | 0.92 ± 0.054 |
| University Hospital of North Tees | 0.99 ± 0.016 | 0.99 ± 0.019 | 0.99 ± 0.015 | 0.99 ± 0.017 | 0.99 ± 0.017 | 0.99 ± 0.018 | 0.99 ± 0.014 |
| University<br>Hospitals of<br>Coventry and<br>Warwickshire | $0.95 \pm 0.009$ | $0.95 \pm 0.011$ | $0.95 \pm 0.011$ | $0.95 \pm 0.014$ | $0.96 \pm 0.017$ | $0.96 \pm 0.014$ | $0.95 \pm 0.014$ |

### EXTERNAL TEST PERFORMANCE

On the external test from Southampton Hospital, consisting of 1,768 WSIs, our model with site-specific standardisation achieved an average accuracy of 94% ± 2.2%, an average sensitivity of 93% ± 1.4%, an average specificity of 95% ± 3.3% and an average AUC of 0.97 ± 0.008 across the five experiments. Performance was consistent with that observed in leave-one-hospital-out cross-validation

### EFFECT OF SITE-SPECIFIC STANDARDISATION

On the training dataset, assessment using leave-one-hospital-out cross-validation showed that site-specific standardisation resulted in a 20-percentage-points increase in accuracy, 40-percentage-points increase in mean specificity and an increase of 0.07 for AUC, while sensitivity was preserved (Table 2).

To determine how much target-site data were required for site-specific standardisation, we repeated the analysis while estimating the site-specific mean and standard deviation from progressively smaller subsets of unlabelled target-site biopsy WSIs. Performance plateaued with as few as 5% target-site biopsy WSIs, with no meaningful improvement when additional biopsy WSIs were used (Supplementary Figure 1).

### RESIDUAL SITE INFORMATION IN LEARNED REPRESENTATIONS

UMAP projections of the biopsy-level representations showed marked differences in site-related clustering according to the standardisation strategy (Figure 2). With global standardisation, representations clustered predominantly according to hospital of origin, with limited overlap between hospitals. Site-specific standardisation substantially reduced this separation and increased overlap between hospitals. Neither standardisation strategy resulted in clear separation between normal and neoplastic biopsy WSIs in the two-dimensional UMAP projections.

**Figure 1.**
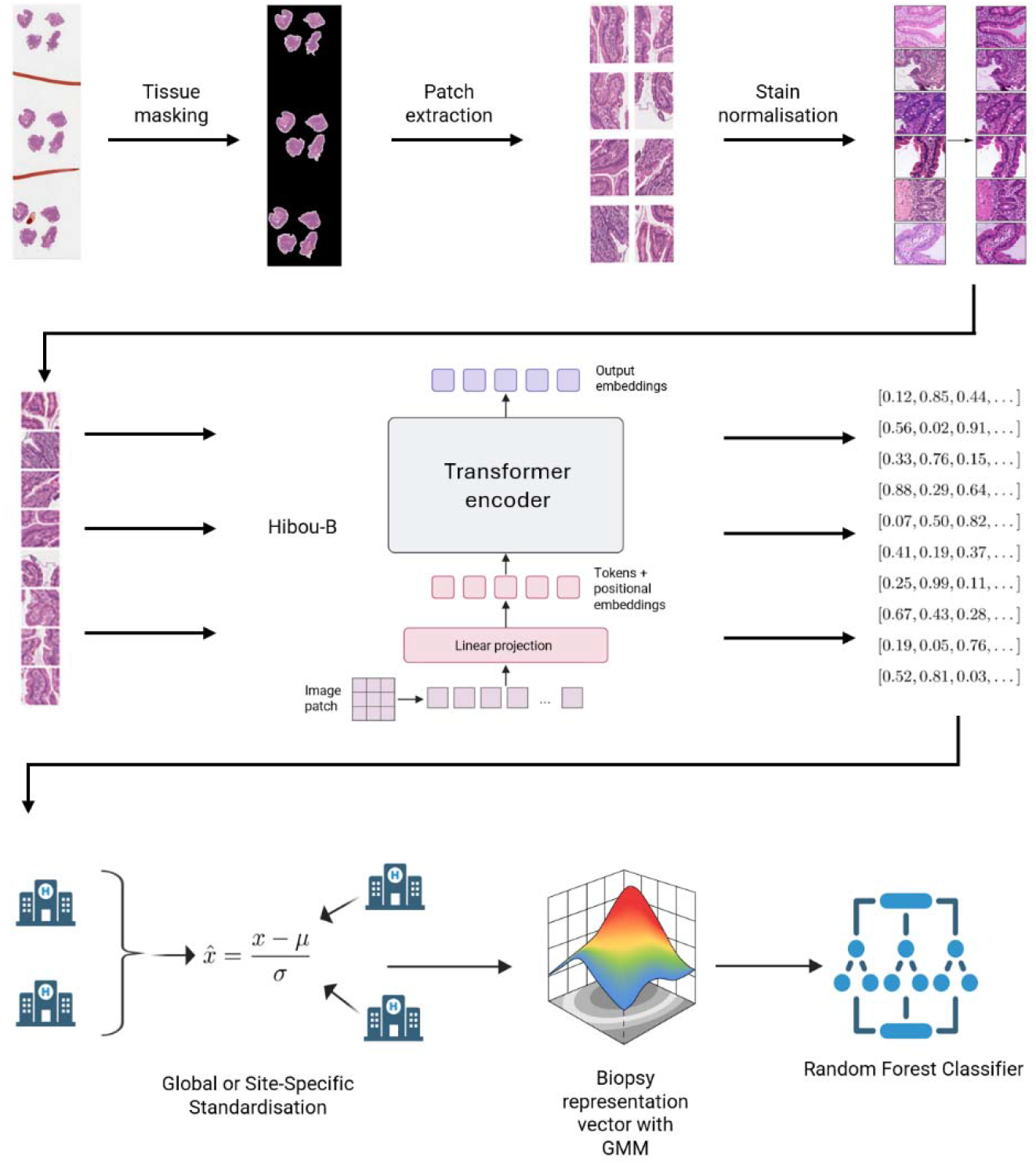
Tissue regions were identified from H&E-stained whole-slide images (WSIs), divided into image patches, and stain-normalised. Patch-level embeddings were extracted using the Hibou-B pathology foundation model. Embeddings were then standardised using site-specific statistics and reduced to 50 dimensions using principal component analysis (PCA). A Gaussian mixture model (GMM) was used to construct a biopsy-level representation by combining mean component-assignment probabilities with mean residual vectors. The resulting 100-dimensional biopsy representation was passed to a Random Forest classifier to classify each biopsy as normal or neoplastic.

**Figure 2.**
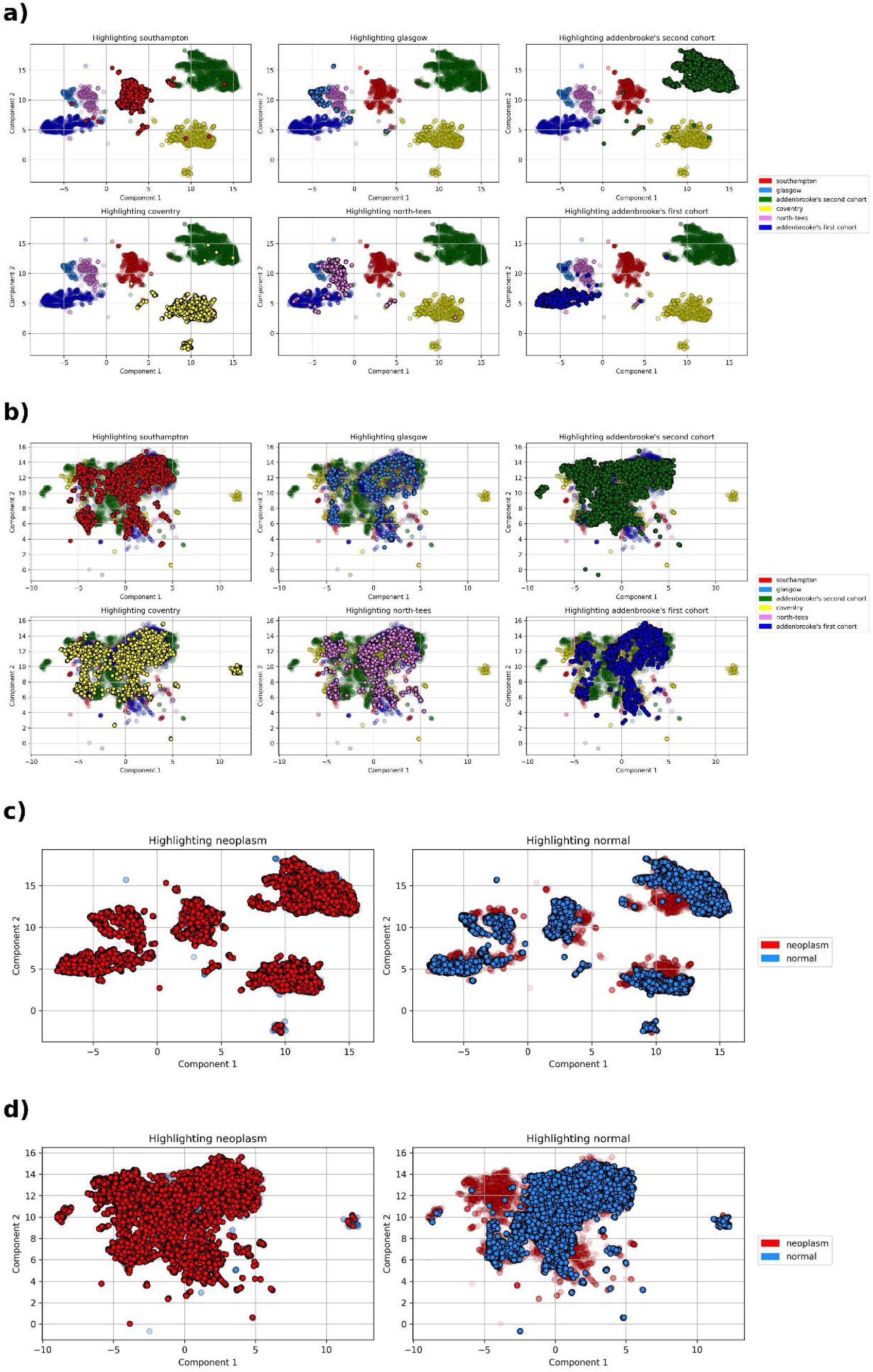
UMAP projections of biopsy-level representations obtained using site-specific and global standardisation. All panels show all 401 neoplastic biopsies and an equal number of randomly sampled normal biopsies. Each point represents a biopsy-level embedding vector generated using PCA and the GMM-based pipeline. The two Addenbrooke’s Hospital cohorts (see Methods) were acquired in different years using different scanners. **a)** Representations obtained using global standardisation, coloured by hospital of origin. **b)** Representations obtained using site-specific standardisation, coloured by hospital of origin. **c)** Representations obtained using global standardisation, coloured by diagnostic label (normal or neoplastic). **d)** Representations obtained using site-specific standardisation, coloured by diagnostic label.

To quantify the residual site information observed in the UMAP projections, we assessed whether hospital of origin could be predicted from individual patch representations. Hospital prediction accuracy decreased from 98% ± 0% with global standardisation to 84% ± 2.5% with site-specific standardisation, indicating that site-specific standardisation reduced, but did not eliminate, site-specific information encoded in the representations.

### COMPARISON WITH MEAN POOLING

Our mean pooling baseline had a better average accuracy (78% ± 9.5%) and specificity (64% ± 24.1%) than our GMM-based pipeline that used global standardisation (Table 2), but it performed less well in every reported metric when compared to the GMM-based pipeline with site-specific standardisation.

## Discussion

In this multicentre study, we developed a ML pipeline for detecting duodenal epithelial neoplasia from H&E-stained biopsy WSIs. In leave-one-hospital-out cross-validation across four development hospitals, our GMM-based pipeline with site-specific standardisation achieved a mean accuracy of 90% and AUC of 0.96, with consistently high sensitivity across hold-out hospitals. Performance was maintained on the independent external cohort from the University Hospital of Southampton, where the model achieved an accuracy of 94%, sensitivity of 93%, specificity of 95%, and AUC of 0.97. These findings demonstrate encouraging cross-site performance despite differences in staining and scanning protocols between participating centres.

The GMM-based representation outperformed simple mean pooling, suggesting that modelling the distribution of patch embeddings within each biopsy may capture information that is lost when embeddings are represented solely by their mean. A key finding was the effect of site-specific standardisation on cross-centre performance.

Compared with global standardisation, site-specific standardisation increased mean cross-validation accuracy by 20 percentage points and specificity by 40 percentage points, while preserving sensitivity. Following site-specific standardisation, UMAP visualisation showed substantially less site-associated clustering, and hospital-of-origin prediction accuracy decreased from 98% to 84%. Together, these findings suggest that site-specific standardisation reduces, although does not eliminate, site-specific information within the learned representations.

Importantly, site-specific standardisation is unsupervised and does not require diagnostic labels from the target institution. In our analysis, performance plateaued when embedding statistics were estimated from as few as 5% of the total sampled unlabelled target-site biopsy WSIs. This suggests that adaptation to a new centre may require only a relatively small set of routine, unlabelled biopsy WSIs.

### Limitations

This study has several limitations. Firstly, the number of biopsy WSIs representing individual types of neoplasm was relatively small, particularly for carcinoma and NET, restricting the accuracy of neoplasm-type-specific performance estimates. Additionally, although we used biopsy WSIs from five different hospitals, one contributed approximately 57% of all biopsy WSIs used in training. Site-specific standardisation also requires access to unlabelled biopsy WSIs from the target institution before deployment. Although performance plateaued with as few as 5% of the total sampled target-site biopsy WSIs in our experiments, this requirement should be evaluated prospectively across additional centres. Finally, external validation was performed using data from a single independent hospital, and further evaluation across additional centres, scanners and staining protocols is required to establish the robustness and generalisability of our approach.

## Conclusions

To our knowledge, this is the first ML pipeline for detecting duodenal epithelial neoplasia in H&E-stained biopsies. Using foundation-model-derived patch embeddings, our GMM-based approach achieved an AUC of 0.96 in leave-one-hospital-out cross-validation across data from the four hospitals in our training set, and maintained an AUC of 0.97 on an independent test set from a fifth hospital. Site-specific standardisation substantially improved cross-centre performance, compared with global standardisation, and required only a small number of unlabelled target-site biopsy WSIs to estimate the necessary embedding statistics. These findings demonstrate the feasibility of a data-efficient approach to ML-based detection of duodenal epithelial neoplasia, and support further evaluation in larger independent multicentre cohorts.

## Data Availability

The data underlying this study are not publicly available because the ethics approval and participant consent agreements do not permit data sharing. The model weights in the present study are available upon reasonable request to the authors

## Contribution

Conceptualization: all authors; Methodology: FDL, FJ; Software: FDL; Validation: FDL; Formal analysis: FDL; Investigation: FDL; Resources: EJS, FJ; Data curation: all authors; Writing – original draft: FDL, FJ; Writing – review & editing: all authors; Visualization: FDL; Supervision: EJS, FJ; Project administration: FJ; Funding acquisition - EJS, FJ.

## Supplementary Materials

### Supplementary Materials A: Number of biopsies needed to estimate parameters for site-specific standardisation

We studied how many biopsy WSIs are needed for site-specific standardisation to perform effectively in an external hospital. We used leave-one-hospital-out cross-validation using our four training hospitals to train and validate our GMM-based pipeline with site-specific standardisation. For each hospital, we used all available neoplastic biopsy WSIs and sampled an equal number of normal biopsy WSIs. We sampled 100 patches from each biopsy WSI. To estimate the parameters for site-specific standardisation for the validation set (mean and standard deviation), we used increasing percentages of the sampled biopsies from the validation set. We repeated each experiment five times.

**Supplementary Figure 1.**
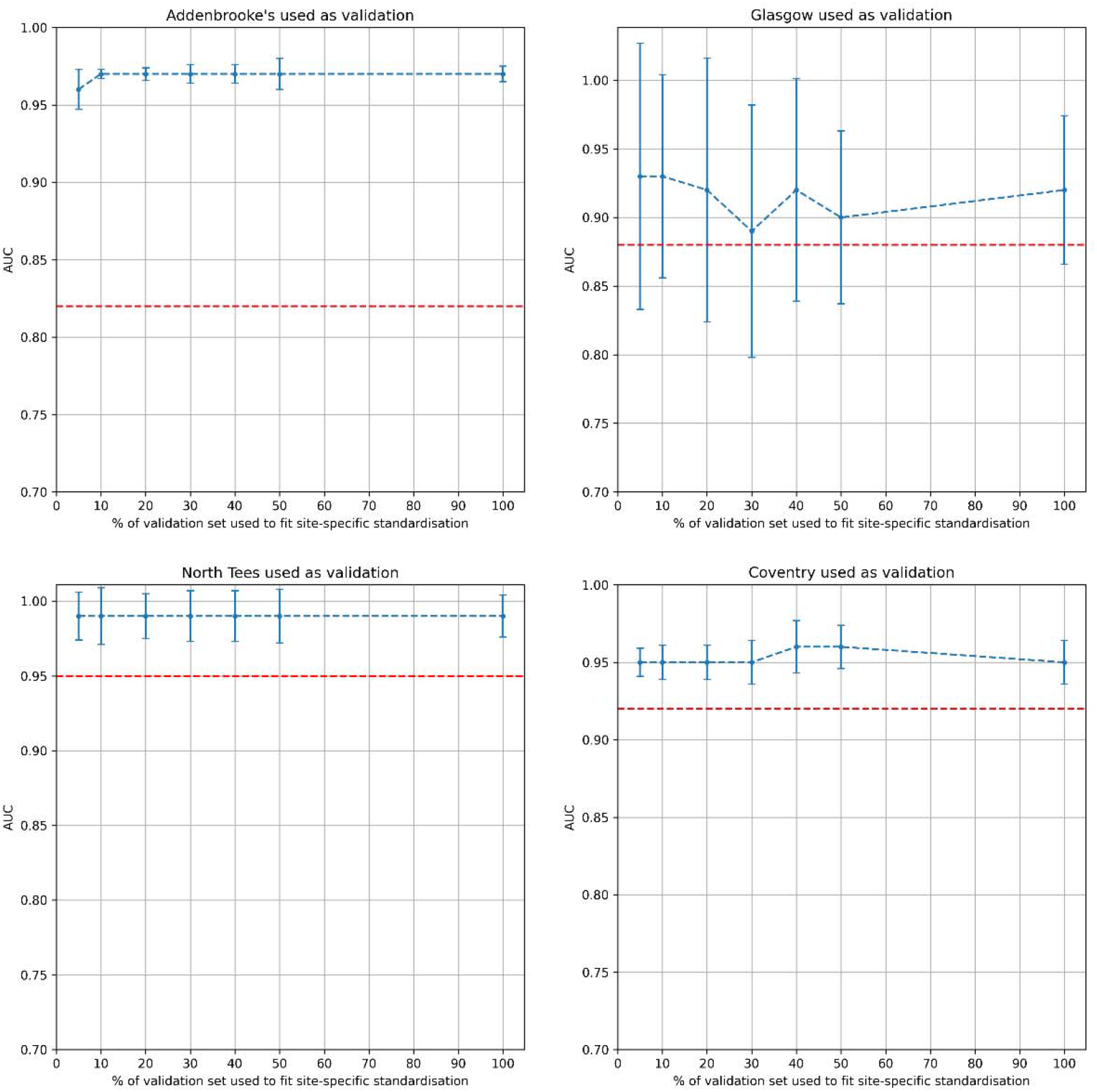
AUC of our GMM using site-specific standardisation in leave one hospital-out cross-validation. Each panel corresponds to one of the four hospitals used as the validation in leave-one-hospital-out cross-validation. The x axis shows the percentage of sampled biopsies used to estimate the parameters of site-specific standardisation. The AUC of our GMM using global standardisation is reported as a dashed red line for reference. The error bars represent the standard deviations, and the points represent the mean across the five experiments.

### Supplementary Materials B: Patient sex and age prediction from patch embeddings

We studied whether our GMM-based pipeline using global standardisation and our mean pooling baseline could predict the sex of our patients (M or F). For this test, in order to remove the variability stemming from the hospital source and the diagnosis, we used a total of 800 Normal biopsies (400 M and 400 F) from Addenbrooke’s Hospital. We randomly sampled 100 patch embeddings for each biopsy. We repeated each experiment five times, each time sampling different biopsies and different patches. Using the same cohort, we assessed whether our pipeline was able to predict the patients’ ages. Our GMM-based pipeline achieved an average accuracy of 59% ± 0.3% when predicting patient sex and an average R^2^ score of 0.11± 0.031 when predicting patient age, while our mean pooling baseline achieved an average accuracy of 58% ± 1.1% when predicting patient sex and an average R^2^ score of 0.14± 0.031 when predicting patient age. Overall, the low performance for both sex and age prediction suggests that these variables are unlikely to be substantial confounders for our pipeline.

### Supplementary Materials C: Classification of type of neoplasm

To assess which type of neoplasm was the most challenging to classify, we trained our GMM with site-specific standardisation using all available neoplastic biopsy WSIs from our four training hospitals and an equal number of normal biopsies sampled from each one of them. We sampled 100 patches from each biopsy. We then tested it on the independent external hospital using all available patch embeddings for each biopsy, as described in the Methods section. We then calculated the AUC for normal vs. adenoma: 0.96 ± 0.014, normal vs. carcinoma: 0.99 ± 0.005, and normal vs. NET: 0.98 ± 0.011. We repeated this experiment five times.

### Supplementary Materials D: Using all available patch embeddings for each biopsy vs. sampling 100 patch embeddings

An ablation study was conducted to evaluate the effect of the patch sampling strategy employed. We compared training our models using all available patch embeddings per biopsy to randomly sampling 100 patch embeddings per biopsy. Due to memory constraints, our experiments were limited to a balanced subset of the training data, consisting of 147 neoplastic biopsy WSIs (50 from Addenbrooke’s Hospital, Cambridge: 50 from the University Hospitals of Coventry and Warwickshire; 33 from the University Hospital of North Tees; and 14 from Queen Elizabeth Hospital, Glasgow), with an equal number of normal biopsy WSIs randomly selected from each respective hospital. Both the GMM-based pipeline with global standardisation (Table 4) and the mean pooling baseline (Table 5) were assessed using the leave-one-hospital-out strategy described above, with each experiment repeated five times. To ensure a fair comparison, both sampling strategies were applied to the same sampled biopsies within each repetition. After having observed that the results between the two approaches were comparable, we opted for using 100 randomly sampled patch embeddings for each biopsy during training in the subsequent experiments, which allowed us to use our computational resources more efficiently. Using all available patch embeddings for each biopsy did not result in a better performance on leave-one-hospital-out validation than randomly sampling 100 patch embeddings for each biopsy when testing the GMM-based pipeline using global standardisation (Table 4). However, the mean pooling baseline showed an increase of 9% in accuracy (Table 5).

**Table 4.** GMM-based pipeline (global standardisation), sampling vs all patches. We report the average accuracy and sensitivity across the 5 repetitions of the GMM-based pipeline using global standardisation on leave-one-hospital-out validation when using 100 randomly sampled patch embeddings for each biopsy or all available patch embeddings. Results are reported as mean ± standard deviation.

| Hospital used as validation | Accuracy (all patches) | Accuracy (100 patches) | Sensitivity (all patches) | Sensitivity (100 patches) |
| --- | --- | --- | --- | --- |
| Addenbrooke's Hospital, Cambridge | 61% ± 3.2% | 58% ± 1.9% | 98% ± 3.5% | 96% ± 3.9% |
| Queen Elizabeth Hospital, Glasgow | 71% ± 12.7% | 71% ± 13.5% | 100% ± 0% | 100% ± 0% |
| University Hospital of North Tees | 63% ± 12.1% | 69% ± 12.4% | 98% ± 2.7% | 98% ± 2.7% |
| University Hospitals of Coventry and Warwickshire | 78% ± 10.8% | 80% ± 8.5% | 83% ± 12% | 95% ± 2.7% |
| Mean | 68% ± 7.8% | 70% ± 9% | 95% ± 7.9% | 97% ± 2.2% |

**Table 5.** Mean pooling baseline with global standardisation, sampling vs all patches. We report the average accuracy and sensitivity across the 5 repetitions of the mean pooling baseline using global standardisation on leave-one-hospital-out validation when using 100 randomly sampled patch embeddings for each biopsy or all available patch embeddings. Results are reported as mean ± standard deviation.

| Hospital used as validation | Accuracy (all patches) | Accuracy (100 patches) | Sensitivity (all patches) | Sensitivity (100 patches) |
| --- | --- | --- | --- | --- |
| Addenbrooke's Hospital, Cambridge | 83% ± 8.5% | 64% ± 1.9% | 94% ± 2.8% | 96% ± 0.7% |
| Queen Elizabeth Hospital, Glasgow | 86% ± 7.8% | 78% ± 7.1% | 100% ± 0% | 100% ± 0% |
| University Hospital of North Tees | 86% ± 1.1% | 87% ± 1.3% | 86% ± 6.2% | 83% ± 5.4% |
| University Hospitals of Coventry and Warwickshire | 87% ± 5.5% | 80% ± 2.9% | 88% ± 2.6% | 82% ± 2.3% |
| Mean | 86% ± 1.7% | 77% ± 9.6% | 92% ± 6.3% | 90% ± 9.1% |

### Supplementary Materials E: Patient-level classification

We trained our GMM-based pipeline using site-specific standardisation using data from our four training hospitals. We used all available neoplastic biopsy WSIs and sampled an equal number of normal biopsy WSIs from each of them. We used 100 patch embeddings from each biopsy during training. We then tested our pipeline on our external test set using all available neoplastic biopsy WSIs and sampling an equal number of normal biopsies. For testing, we used all available patch embeddings for each biopsy.

Some patients had more than one WSI of their biopsy, which arose from the cutting of multiple sections of the biopsy tissue onto separate glass slides, which were then scanned. To perform classification at the patient level, we assigned the patient biopsy WSI ground-truth label as neoplastic, if at least one of their biopsy WSIs was labelled as neoplastic; otherwise, the patient was labelled as normal. Similarly, a patient biopsy WSI was predicted to be neoplastic if at least one of their biopsies was predicted as neoplastic by our pipeline; otherwise, the patient biopsy WSI was predicted as normal. We repeated this experiment five times.

Using our GMM-based pipeline using site-specific standardisation on our test set, we obtained a mean accuracy of 0.93 ± 0.023, a mean AUC of 0.95 ± 0.014, a mean sensitivity of 0.91 ± 0.013 and a mean specificity of 0.95 ± 0.034.

